# Interpretable-by-design Deep Survival Analysis for Disease Progression Modeling

**DOI:** 10.1101/2024.07.11.24310270

**Authors:** Julius Gervelmeyer, Sarah Müller, Kerol Djoumessi, David Merle, Simon J. Clark, Lisa Koch, Philipp Berens

**Affiliations:** Hertie Institute for AI in Brain Health, University of Tübingen, Tübingen, Germany; Department of Ophthalmology, University Eye Clinic, University of Tübingen, Tübingen, Germany; Department of Diabetes, Endocrinology, Nutritional Medicine and Metabolism UDEM, Inselspital, Bern University Hospital, University of Bern, Bern, Switzerland

**Keywords:** Interpretability, Deep survival analysis, Disease prognosis

## Abstract

In the elderly, degenerative diseases often develop differently over time for individual patients. For optimal treatment, physicians and patients would like to know how much time is left for them until symptoms reach a certain stage. However, compared to simple disease detection tasks, disease progression modeling has received much less attention. In addition, most existing models are black-box models which provide little insight into the mechanisms driving the prediction. Here, we introduce an interpretable-by-design survival model to predict the progression of age-related macular degeneration (AMD) from fundus images. Our model not only achieves state-of-the-art prediction performance compared to black-box models but also provides a sparse map of local evidence of AMD progression for individual patients. Our evidence map faithfully reflects the decision-making process of the model in contrast to widely used post-hoc saliency methods. Furthermore, we show that the identified regions mostly align with established clinical AMD progression markers. We believe that our method may help to inform treatment decisions and may lead to better insights into imaging biomarkers indicative of disease progression. The project’s code is available at github.com/berenslab/interpretable-deep-survival-analysis.

## 1 Introduction

Age-related macular degeneration (AMD) is the main cause of legal blindness in developed countries, caused by cumulative damage to the central retina leading to loss of central vision. This progressive disease severely impacts patients’ quality of life by impairing tasks requiring sharp vision, motivating the need for early detection and intervention. AMD is characterised by retinal changes, such as drusen and pigment abnormalities, and is typically classified into early, intermediate and late disease stages, where vision is mostly endangered in the latter [12]. Retinal fundus images can reveal such indicators of AMD and consequently, multiple studies have developed deep learning solutions to accurately detect the disease stage [13,19] or predict the conversion to late AMD by a specified time [4,25]. However, these black-box models do not provide inherent interpretability and could only be explained by post-hoc saliency maps. These, however, lack reliability, which is particularly problematic in high-risk medical applications [3,22,21]. As an alternative, interpretable-by-design models such as the Sparse BagNet [10] have recently been proposed for classification tasks on retinal images. In this model architecture, evidence is gathered locally, represented in explicit evidence maps, which is then aggregated to predict the overall class. The class evidence maps can be visualised as heatmaps overlaid on the input image, highlighting the contribution of small local regions to the final prediction. However, the Sparse BagNet was developed for a classification setting and had not yet been adapted for disease progression modeling.

In this paper, we developed an interpretable-by-design model for AMD disease progression using a survival analysis framework. Survival analysis models are a popular choice for time-to-event modeling which predict for each individual the probability to “survive” – i.e., to *not* observe – an event of interest within a time frame, such as progressing from early to late AMD. Classical survival analysis uses simple linear models such as the Cox proportional hazards (CoxPH) model [8], but deep survival models have been proposed that parameterise CoxPH models with neural networks [11,16,23]. In this work, we first integrated the current state-of-the-art model architectures for AMD progression modeling [4,25] into a deep survival model. We then replaced the backbone neural network with a Sparse BagNet, which provides an inherently interpretable evidence map for the predicted survival curve of an image, i.e. the risk of disease progression. To the best of our knowledge, this is the first image-based interpretable-by-design deep survival analysis model. We provide an overview over our method in Fig. 1.

**Fig. 1.**
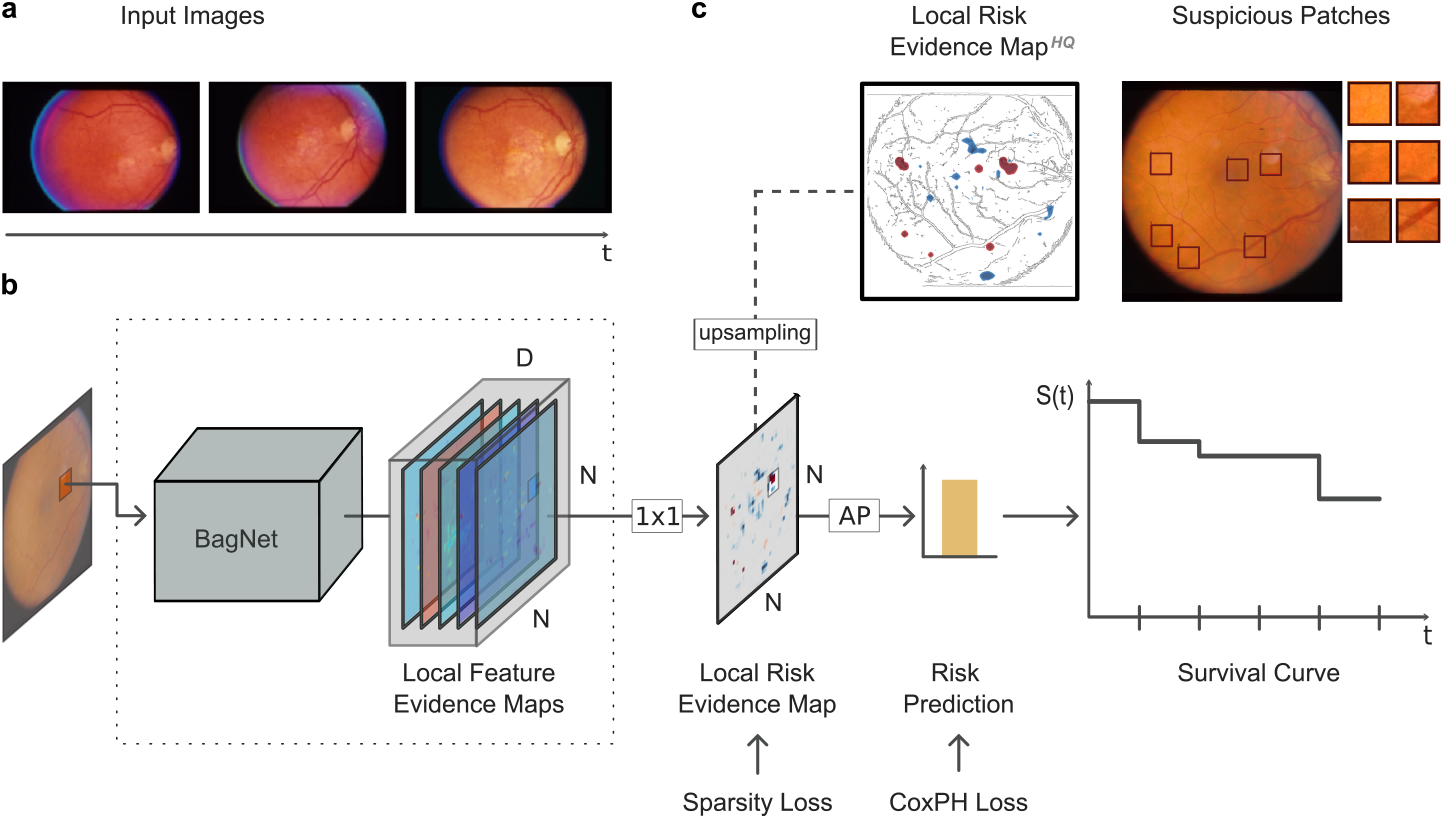
Interpretable-by-design Deep Survival Model for AMD Progression. **a**. Fundus images of an eye at various screenings. **b**. An image is fed into the BagNet survival model, which first yields local feature evidence maps (D the features dimension, N its width and height) as in [6]. After a 1 *×* 1 convolution, we receive a local risk evidence map, on which we apply a sparsity loss. Average pooling (AP) yields the final risk prediction as in [10] on which we apply a CoxPH loss. In a second step, the prediction is translated into a survival curve *S*(*t*), the individual’s probability of “surviving” the event of interest – conversion to late AMD. **c**. The local risk evidence map allows intuitive interpretation of predictions (left). Clinicians could be provided with suspicious patches, i.e., regions that the model found to indicate a conversion risk (right).

## 2 Methods

### 2.1 Dataset and Preprocessing

We worked with data from the Age-Related Eye Disease Study (AREDS), a longitudinal study sponsored by the National Eye Institute, USA [1] which is available upon request^1^ for research purposes. A total of 4,757 participants between 55 and 80 years of age were screened over a course of 12 years to study the natural progression of age-related eye diseases. The study was approved by the institutional review boards at all participating clinical sites and participants gave written consent [2]. We filtered out fundus images that were missing information such as the AMD severity score (1-9: increasing AMD severity; 10-12: central geographic atrophy and/or neovascular AMD, i.e., late AMD) and used the remaining 133,293 macula-centered fundus images for this project. Images came in pairs of photographs of the same fundus from slightly different angles to allow retinal specialists for depth impressions. We randomly selected one of the two views from each pair and excluded images after conversion to late AMD. Images were first resized to a height of 350 pixels and then cropped to a width of 350 pixels from the center. We further applied random resized cropping, flipping, color jitter, and rotation with the settings from [15], as these proved useful to enhance retinal disease detection. We split data into 60% training, 20% validation, and 20% test set, keeping a participant’s records in the same split.

We defined the label targets *event* and *time* as follows: the event was 1 if the eye’s AMD severity score reached 10 (late AMD) any time during the study and 0 otherwise (no late AMD). For this project, we defined time as the relative duration from an eye scan to the screening after which the eye was first diagnosed with late AMD or, if there was no such diagnosis, the time until the patient’s last screening session. For time-dependent evaluation, we defined a “case” as an eye that converted to late AMD before or at that time.

### 2.2 Interpretable-by-design Deep Survival Model

#### Deep Survival Analysis

In survival analysis, CoxPH models provide a scalar output that can be used in a second step to estimate a set of risk predictions over time. We denote

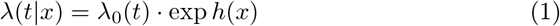

as the hazard function, where *λ*_0_(*t*) is the population wide baseline hazard function and *h*(*x*) the individual log partial hazard based on a set of features *x*. A hazard at time *t* is the instantaneous risk to observe an event of interest at (an infinitesimally small interval around) *t*. Classically, *h*(*x*) is a linear function. For deep survival analysis, it can be parameterised by a neural network with any choice of architecture (ResNet-50, Inception-v3 or others) with one output node: we can apply a sigmoid function to the model’s output logit and interpret the result as an estimate of the log partial hazard 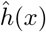 . Here, this refers to the subject’s relative risk to develop late AMD. This framework allows to directly work on fundus images as inputs *x* and to train end-to-end. We estimated the baseline hazard function *λ*_0_(*t*) using Breslow’s maximum likelihood approach [7] given the training data. The survival curve is given by

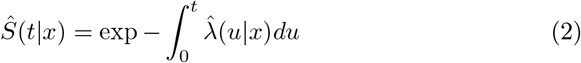

which represents the probabilities not to convert to late AMD up to time *t*.

#### Adding Inherent Interpretability

Instead of using a non-interpretable standard architecture to parameterise *h*(*x*), we used a modified inherently interpretable model, the bag-of-local-features model, referred to as BagNet [6]. The BagNet is based on the ResNet-50 architecture, but changes in strides and the kernel sizes restrict the model to work on local image patches of size *q × q*. After slight modification, the BagNet architecture can yield an explicit and local map of activations for each class: one pixel of each final feature map represents the local class evidence from an image patch in the input image [10]. Note that standard ResNets learn potentially global features and their interactions, while the BagNet learns the local evidence in an image patch. This eliminates the need for post-hoc saliency maps or the post-hoc analysis of convolutional filters. For the BagNet, we chose a receptive field size of *q* = 33 pixels and, following [10], we applied a sparsity constraint to the loss to avoid cluttered evidence maps (see Suppl. Fig. 1 for the selection of the sparsity coefficient). The model decision is then obtained by spatial average pooling, resulting in a final risk prediction logit. Correspondingly, the Sparse BagNet for survival analysis produces one class evidence map for the risk of disease progression and, as a result, allows a direct and intuitive interpretation of the survival predictions. In contrast, the baseline models are classification models that are trained once for each queried prediction time, resulting in a set of models, each of which has a saliency map to consider.

#### Implementation Details

We initialised the model with pre-trained weights from ImageNet and trained it for up to 50 epochs on the training set with a batch size of eight. Weights were updated based on the CoxPH loss [11,16], that is, the negative log likelihood loss of the log partial hazard *ĥ*, adapted from the auton-survival package [18]. We applied Breslow’s method as implemented in scikit-survival [20]. We used the Adam optimiser at a learning rate of 1.6*e*–5, as determined by a hyperparameter search. Training was conducted using a NVIDIA GeForce RTX 2080 Ti GPU and the PyTorch framework.

### 2.3 Baselines

Recently proposed end-to-end trained AMD progression models consist of one classification model for each queried time point [4,25,26]. As our baselines, we implemented classification models from Yan et al. [25] and Babenko et al. [4] that, similar to our model, are based only on fundus images. Both studies utilise Inception-v3 architectures to predict whether an eye converts to late AMD. However, Yan et al. use one fundus image per eye and screening as input, while Babenko et al. use both images of a stereo pair as inputs to separate Inception-v3 modules with shared weights and average their predictions after the activation function. Further, Yan et al. use the Adam optimiser and Babenko et al. use stochastic gradient descent. We re-implemented both models in our framework as close as possible to the originally reported settings. Training was conducted at a learning rate of 1*e*–4, with the number of epochs, early stopping, batch size, image size and data augmentation set according to our proposed model. We used a binary cross-entropy loss and set the event label to 1 if the eye was first diagnosed with late AMD at or before the inquired year. If an eye did not convert to late AMD and the subject’s last screening was before the inquired year, we could not extract classification labels and therefore had to exclude these records.

### 2.4 Evaluation Strategy

We evaluated the disease progression models using the area under the receiver operating characteristic curve (AUROC), Brier Score, and area under the precision-recall curve (AUPRC), adjusted for time-dependent predictions [17]. We chose the cumulative sensitivity/dynamic specificity approach to calculate AUROC using the scikit-survival package, which assesses the discrimination of eyes at higher risk from eyes at lower risk. Here, *cumulative cases* are subjects who have experienced the event up to a given time, while all eyes that have not (yet) experienced the event are referred to as *dynamic controls*. We computed Brier Scores as a measure of model calibration as implemented in scikit-survival and AUPRC as a performance measure focusing on cases using the R package time-ROC [5]. We additionally provide the metrics variants that are not adjusted for time-dependence in Suppl. Tab. 1. During training, we evaluated each epoch based on the Integrated Brier Score (IBS) of the survival probability predictions on the validation set. Training was stopped if the IBS did not improve within ten consecutive epochs. We selected the final model weights based on the epoch with the lowest IBS.

## 3 Results

We developed an architecture for disease progression modeling that achieves inbuilt interpretability through an explicit evidence map layer that contains the local evidence for disease progression. We applied the model to the prognosis of AMD progression and predicted the probability of not converting to late AMD within one to five years and compared it to state-of-the-art models.

### 3.1 Interpretable-by-design Model Achieves SOTA Performance

Our model performed comparably to state-of-the-art AMD progression models (Tab. 1) and only slightly worse in terms of AUROC and Brier Scores. We studied which of the components of our model were responsible for the performance difference compared to the baseline models. We found that training the Sparse BagNet as classification models reduced the performance, while replacing the Sparse BagNet with an unmodified ResNet-50 improved it (see Suppl. Fig. 2). This indicates that the CoxPH model training was not responsible for the slightly decreased performance, but rather the BagNet architecture.

**Table 1.**
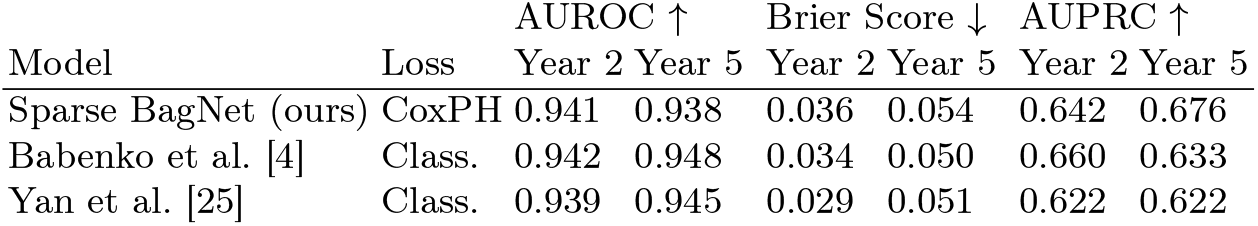
Performance of our interpretable-by-design model compared to black-box SOTA models on the unseen test data (see Methods for details).

| Model | Loss | AUROC ↑ | | Brier Score ↓ | | AUPRC ↑ | |
| --- | --- | --- | --- | --- | --- | --- | --- |
|  |  | Year 2 | Year 5 | Year 2 | Year 5 | Year 2 | Year 5 |
| Sparse BagNet (ours) | CoxPH | 0.941 | 0.938 | 0.036 | 0.054 | 0.642 | 0.676 |
| Babenko et al. [4] | Class. | 0.942 | 0.948 | 0.034 | 0.050 | 0.660 | 0.633 |
| Yan et al. [25] | Class. | 0.939 | 0.945 | 0.029 | 0.051 | 0.622 | 0.622 |

### 3.2 Heatmaps Provide Faithful and Intuitive Model Interpretation

We extracted the evidence map from our model showing the local risk of conversion to late AMD – this map may show positive entries indicating higher risk of conversion in some regions or negative entries indicating lower risk. Crucially, the final risk prediction is simply formed by spatially averaging this evidence map so that it provides a faithful visualisation of the model’s decision process when overlaid on an image (Fig. 2a). For example, the evidence map overlaid on fundus images of a non-converting participant at three subsequent screenings highlighted mostly regions indicating low risk of conversion (1^st^ row of Fig. 2a) and the corresponding predicted survival curve stayed close to 1 (Fig. 2b). In contrast, evidence maps overlaid on fundus images of participants who converted to late AMD showed evidence for increased predicted risk in sizable portions of the fundus images (2^nd^ and 3^rd^ rows of Fig. 2a). Accordingly, the survival curve showed lower survival probability with conversion events indeed happening within a relatively short time frame of a few years (Fig. 2b). Remarkably, even though the image perspective shifted slightly between imaging time points the highlighted high risk regions were consistently identified (Fig. 2a).

**Fig. 2.**
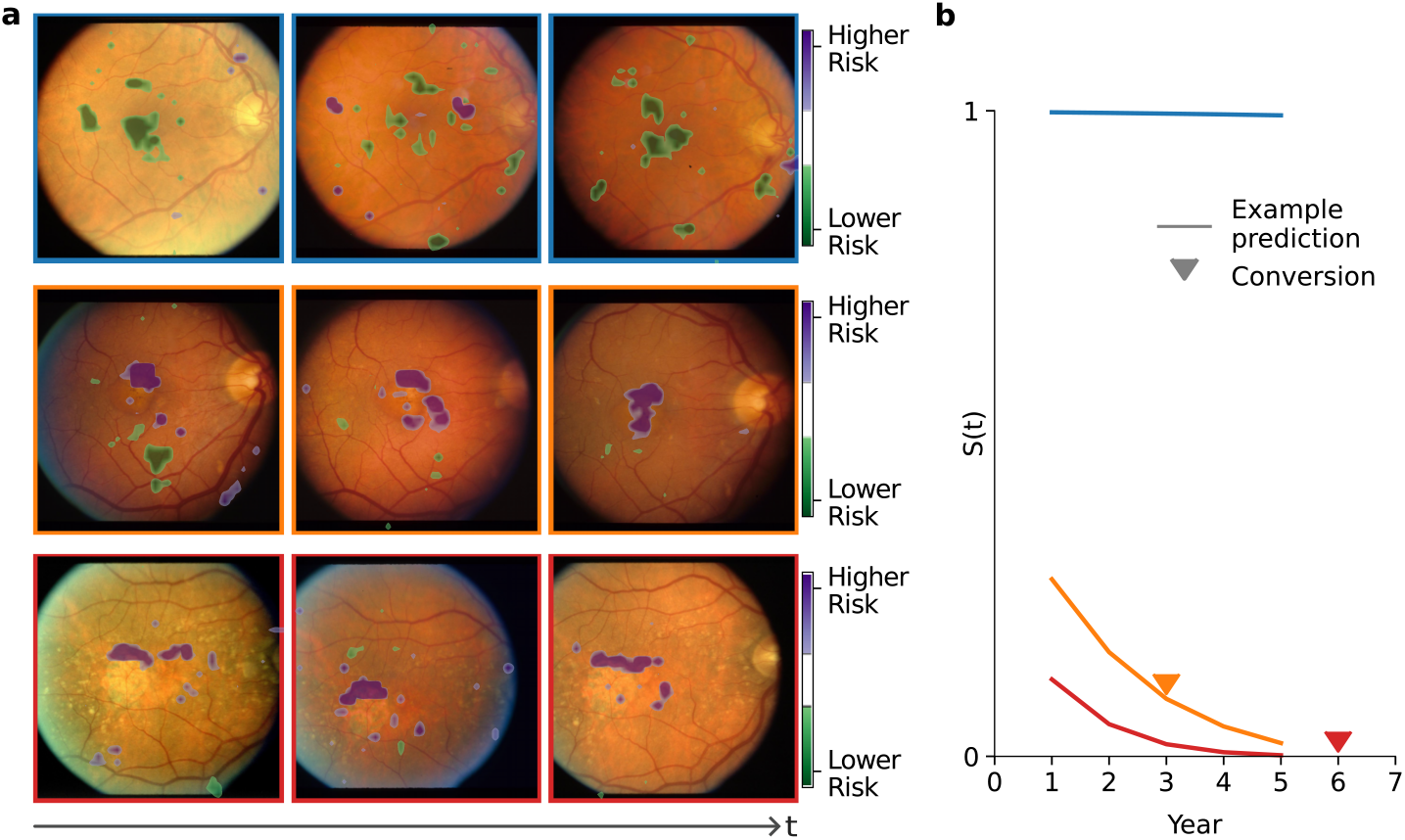
**a**. Examples of predicted local evidence for the risk of AMD conversion for images from multiple consecutive screenings from a healthy eye (blue) and eyes that later converted to late AMD (orange, red). **b**. Example survival curves show predicted probabilities of surviving the event of interest – conversion to late AMD. An arrow indicates the time of late AMD onset.

Based on our evidence maps, the most important image regions can be bounded by boxes sized according to the model’s receptive field and could be shown to clinicians to help them understand the prediction and potentially refine their own assessment (Fig. 3). In contrast, saliency maps for state-of-the-art models obtained using post-hoc gradient based techniques were often much less spatially confined (see Suppl. Fig. 3).

**Fig. 3.**
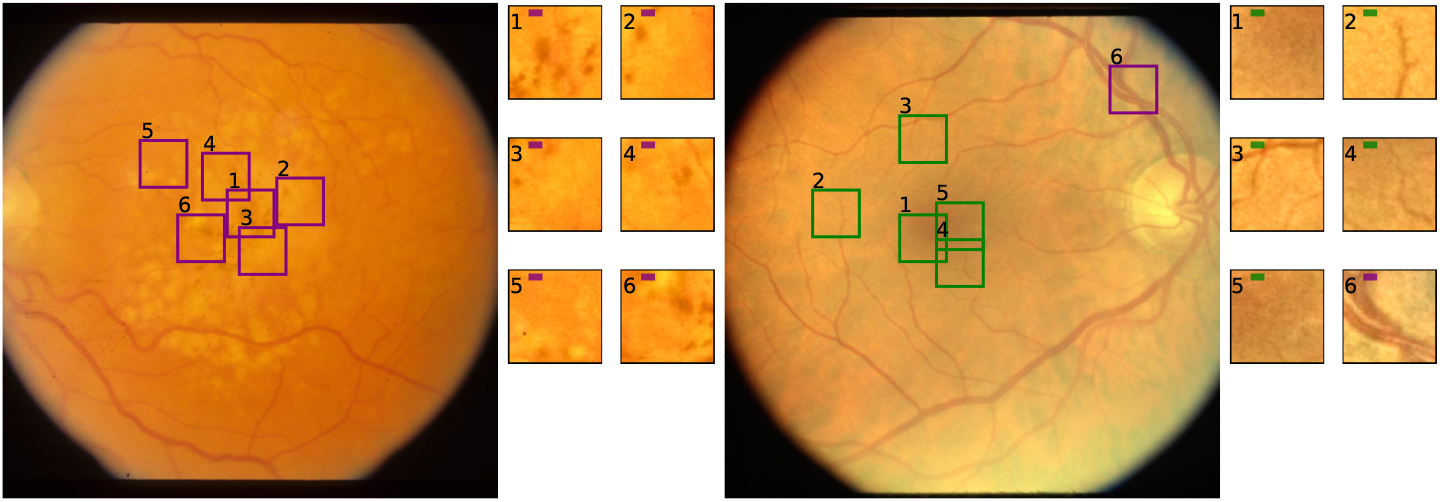
Two examples of interpretable model outputs. Patches show regions with high evidence for a risk to convert to late AMD (purple) and regions that provide evidence for a low AMD conversion risk (green). As the patch label number increases, the importance of the patch decreases.

### 3.3 Heatmaps Capture Regions Known to Indicate AMD Progression

We next analysed to what extent the patches extracted from our evidence maps corresponded to known signs of conversion to late AMD. To this end, DM, a senior resident in Ophthalmology with experience in AMD research, annotated image patches. We included a random selection of the six patches with the highest predicted risk from 20 images from a pool of confident test set predictions for converters (for example Fig. 3). We found that our model focused mostly on regions known to be associated with AMD, with 88.3% of the patches displaying known indications. Most prominently, 38/120 patches showed pigment mottling and 34/120 patches showed soft drusen, both indicative of AMD progression, and 32/120 patches already showed signs of atrophy. The patches shown in Fig. 3 (*left*) for instance contain either soft drusen, pigment mottling or both. In contrast, for non-converters, we could observe that the model focused on unremarkable retinal tissue in the macula or small blood vessels as indicative of low risk (Fig. 3, *right*).

## 4 Discussion

In this work, we introduced the first image-based interpretable-by-design deep survival model for modeling the risk of disease progression and applied it to the risk prediction of conversion to late AMD from fundus images. To this end, we combined a CoxPH survival model with a Sparse BagNet, which yielded highly localised evidence maps faithfully incorporated into the risk prediction process, which is desirable also for ethical reasons [14]. The high-risk areas identified by our model mostly corresponded to established signs of imminent conversion to late AMD. Our model uncovered the AMD risk areas without any prior knowledge from the AREDS dataset alone, indicating the model’s potential for image-level biomarker discovery. In contrast, post-hoc saliency maps computed for state-of-the-art models were much less localised and do not yield faithful reflections on the model’s decision making process [21]. Alternatives to the BagNet backbone include prototype models, which learn prototypical image parts and provide them for interpretability, and deep learning-based additive models such as the EPU-CNN [9], which provides contribution scores for colour and texture concepts along with their spatial relevance. These methods would be worth exploring in the context of disease prognosis. To date, however, prototype models still suffer from imprecise explanations [24], and the EPU-CNN’s interpretations require explicitly computed feature maps, typically based on luminance, color or frequency content. The Sparse BagNet, on the other hand, provides intuitive pixel importance for interpretation, making it well suited for clinical applications and finding novel image-based biomarkers. As the inductive bias of our model fits well to task structures involving small disease features, we expect it to generalise to other medical imaging tasks, including progression modeling for diabetic retinopathy. In summary, our interpretable-by-design deep survival model based on Sparse BagNets opens up new possibilities for trustworthy risk-modeling from medical images beyond ophthalmology and may help to identify new early indications of disease progression which could easily be overlooked by humans.

## Data Availability

Data is available through the dbGaP. Code and model weights are available on Github, see the abstract.

https://github.com/berenslab/interpretable-deep-survival-analysis

https://www.ncbi.nlm.nih.gov/projects/gap/cgi-bin/study.cgi?study\_id=phs000001.v3.p1

## Acknowledgments

This project was supported by the Hertie Foundation, the Else Kröner Medical Scientist Kolleg “ClinbrAIn: Artificial Intelligence for Clinical Brain Research”, the Bundesministerium für Bildung und Forschung through the EYERISK project (BMBF, FKZ 01ZZ2319A) and the German Science Foundation (BE5601/8-1 and the Excellence Cluster 2064 “Machine Learning — New Perspectives for Science”, project number 390727645). The authors thank the AREDS participants, and the AREDS Research Group for their valuable contribution to this research. Funding support for AREDS was provided by the National Eye Institute (N01-EY-0-2127).

## Footnotes

1 https://www.ncbi.nlm.nih.gov/projects/gap/cgi-bin/study.cgi?study_id=phs000001.v3.p1

