## Supplementary File for "Interpretable-by-design Deep Survival Analysis for Disease Progression Modeling"


### Supplementary Material

**Table 1.** Model evaluation using scikit-learn metric implementations: our interpretable deep survival model compared to baselines on the test set. These metrics are not adjusted to time-dependent data.

| | | AUROC ↑ | | Brier Score ↓ | | AUPRC ↑ | |
|---|---|---|---|---|---|---|---|
| Model | Loss | Year 2 | Year 5 | Year 2 | Year 5 | Year 2 | Year 5 |
| Sparse BagNet (ours) | CoxPH | 0.933 | 0.922 | 0.034 | 0.047 | 0.495 | 0.522 |
| Babenko et al. | Class. | 0.936 | 0.927 | 0.028 | 0.044 | 0.564 | 0.542 |
| Yan et al. | Class. | 0.929 | 0.922 | 0.025 | 0.044 | 0.558 | 0.426 |

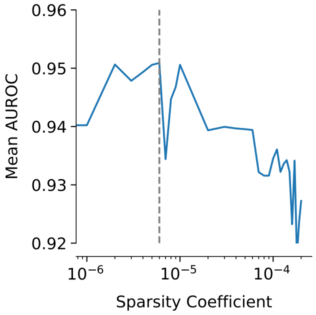


**Fig. 1.** Mean AUROC performance for different values of the sparsity coefficient on the validation set (dashed line shows the selected hyperparameter value of $6e{-}6$).

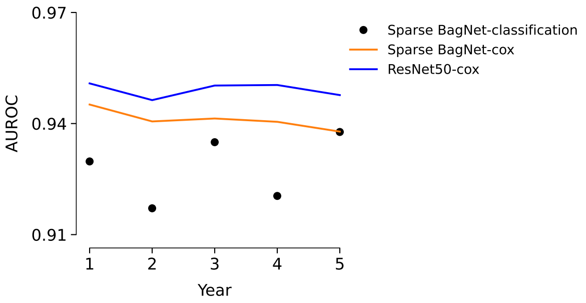


**Fig. 2.** Illustration of the ablation from Sec. 3.1. AUROC performance of our interpretable deep survival model (orange) compared to the same architecture in a classification approach (black) and a ResNet-50 as a CoxPH survival model (blue).

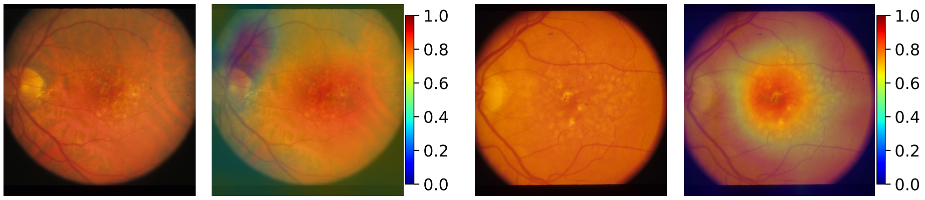


**Fig. 3.** Example visualisations of gradient-based saliency maps from an Inception-v3 model by Babenko et al. using GradCam after the layer block "Mixed 7c".
